# Reform of the intermediate level of the health system in the Democratic Republic of the Congo: Adaptations and limits in the stabilization of the personnel of the Provincial Health Division: A cohort study

**DOI:** 10.64898/2026.06.17.26355888

**Authors:** Charles Ruhangaza Mushagalusa, Daniel Garhalangwanamuntu Mayeri, Amani Ngaboyeka Gaylord, Christine Murhimalika Chimanuka, Pacifique Mwene-Batu, Mwembo Tambwe Albert, Mukalay wa Mukalay Abdon, Bisimwa Balaluka Ghislain

## Abstract

**Background:** Human resources are one of the pillars of health systems. Since the World Health Organization’s report on human resources issues, several countries have integrated this component into the various reforms aimed at strengthening their health systems. This study aims to explore the effects of reforming the intermediate level of a health system operating in a fragile state context.

**Methodology:** Our study was conducted in the Democratic Republic of Congo (DRC). It was a cohort study of the staff of the 14 Provincial Health Divisions (PHD) out of the 26 existing in the DRC. We established a database of the staff of these 14 PHD from 2016, just after the implementation of the intermediate level reform and the allocation of this staff by the Ministry of Health. We did a recall in 2021, in each of these PHD to survey this staff through a structured questionnaire and supplemented by the files of the agents available in each PHD. Sociodemographic, economic and academic variables were collected and analyzed. Data were entered into an Excel 2016 database and processed with SPSS software version 25. The chi-square test was used for comparison of proportions with a statistical significance level of p < 0.05. Risk ratios ratios (RR) and their 95% confidence intervals were calculated as measures of association. The error threshold α was set at 5%.

**Results:** A total of 657 agents with an average age of 45.2 years had been identified in 2016 at the start of the survey and in 2021, 118 or 18% of them were no longer part of the PHD agents. Among the causes of absence noted: 48% of agents placed on leave, 16% promoted to other functions within the health system, 16% desertion and dismissal and 11% cases of death. 19.8% of absentees are executives, 19.5% men against 10.3% women; 22.3% of absentees in unstable provinces against 16.6% in stable ones. The factors associated with the absence of agents in the PHD remain the reaching of retirement age [RR (95% CI) = 5.5 (1.2-24.9)] and male agents [RR (95% CI) = 3.2 (1.3-7.9)]. Among the agents who remained, 92% kept their initial position, 6% were subject to an internal permutation accompanied by a promotion. The factors associated with the stability of human resources at the level of the Provincial Health Division are: female gender, manager with experience or seniority > 5 years, Age > 35 years, Stable province, Presence of a partner bonus.

**Conclusion:** Even in a crisis and fragile context, health system reform is possible. It is possible to organize staff recruitment through a selection process independent of the political authorities of the Ministry of Health and supported by the technical services of the Ministry and partners. Experience and the presence of a financial bonus are motivating factors for staff stability. The involvement of Technical and Financial Support Partners in the recruitment process helped the Ministry of Health to minimize political influence in the recruitment of middle-level executives.

## Introduction

### Context for the implementation of the health care systems

Health sector reforms have become a global phenomenon during the last two decades of the 20th century. Since the World Health Organization’s report on human resources, several countries have integrated this component into various reforms aimed at strengthening their health systems. While each country proceeds in its own way, there is general agreement that radical changes in health care delivery are needed to take full advantage of the opportunities for improving global health [1].

All health system reforms must balance four often contradictory objectives: ensuring the financial viability of the systems, equity of access to care, the quality of care, and finally the freedom and comfort of patients and professionals. [2]. Most reforms can be grouped into three broad categories: financial reform, organizational changes, and policy changes. Organizational reforms are designed to overcome weaknesses in management structures and the lack of performance-based incentives in the public sector. Typical reform measures include decentralizing decision-making levels, encouraging public-private partnerships, and establishing integrated services [3].

According to the WHO World Health Report 2001, most developing countries have initiated efforts to improve their health systems since the late 1980s. These actions have been triggered by a variety of factors: the transition from centrally planned to market economies; inadequate health financing during periods of financial crisis; the inability of many citizens to access basic health services; and the poor quality and inefficiency of existing health services. [5].

In many countries, the lack of good governance had contributed to the low priority given to health development as a component of sustainable human development. Political instability, as well as civil unrest and wars, had also played a major role in the disruption of existing weak health services and in the rise in the incidence of disease and disability [1].

To address these problems, many governments have launched health sector reforms, which take the form of intensive, long-term efforts to strengthen and improve health systems and, ultimately, the health of the nation. [6]. WHO has encouraged low– and middle-income countries (LMICs), including “fragile states,” to implement reforms to improve the performance of their health systems [7,8]. These reforms generally focus on safeguarding the six pillars of the health system as defined by WHO: service delivery, human resources, financing, health information, medicines and equipment, and governance and leadership [9].

### Health system organization in the DRC

The DRC, a Central African country with an estimated population of 90 million inhabitants [10] and an area of 2,345,000 km2. It is the second largest country in Africa after Algeria. The DRC is divided into 26 administrative provinces. The communication routes are deficient (less practicable road network, a few air lines between certain cities and at an irregular rate) thus making contacts between the central administration and the provinces difficult [19]. It is a country classified in the category of “fragile states” due to its instability for nearly 3 decades with long periods of armed conflicts. [25, 26, 27]. The instability remains permanent and is causing several deaths and the displacement of thousands of people, particularly in the eastern part of the country, notably in the provinces of North Kivu, South Kivu, Maniema and Ituri.

For decades, the proportion of the state budget allocated to health has varied between 4 and 6% and more than 70% of health funding comes from development aid [28].

The DRC health system has three levels. The central level (composed of the Minister of Health and his cabinet, the General Secretariat for Health, the General Inspectorate of Health, central directorates and specialized programs) which develops standards and provides broad guidelines for health policy. The intermediate level is made up of 26 PHD and 26 Provincial Health Inspectorates (PHI) corresponding to the 26 administrative provinces. The main mission of the PHD is to provide local supervision of the Health Zones. The operational level is made up of HZ, numbering 519 in total, which ensure the implementation of Primary Health Care (PHC) for the population [20].

In the DRC, the implementation of PHC is decentralized and falls under the jurisdiction of the provinces. and the PHD enjoy relative autonomy of operation.

The DPS operates with six offices (Technical Support, Resource Management, Information and Research, Hygiene, Inspection-Control and Education) under the leadership of a provincial management team, the Provincial Division Head and 6 office heads [20].

### Context and process of health system reform in the DRC

In the DRC, according to the results of an assessment conducted around the 2000s, the health system was described as inadequate [10]. The WHO ranking in its report on world health in 2000 also confirmed this [11]. Indeed, between the end of the 1990s and the beginning of the 2000s, the DRC’s health system was severely impacted by recurring socio-economic and humanitarian crises and showed its inability to meet the health needs of the Congolese population [12].

In order to address these issues, In 2006, the DRC committed to reforming its health system in the midst of the post-conflict period; thus, the Health System Strengthening Strategy (HSSS), developed by Congolese executives, including those from the Ministry of Health, was made public at the end of 2006 and was revised in 2010. One of the axes of this strategy explicitly provides for reforming the intermediate (provincial) level of the health system. The purpose of this reorganization is to support the revitalization of the health district (called Health Zone in the DRC), a priority conditional on the strengthening of the Congolese health system [10,13,14]. The reform of the intermediate level aimed, through its new organizational mode, to respond to two major concerns: i) to take into account the complexity of health systems in terms of epidemiological particularities, service delivery, management of human and financial resources and health information in an environment that is difficult to say the least [15, 16]; ii) be accepted by key stakeholders for implementation [13].

In the DRC, the reform of the intermediate level of the health system has been undertaken since the 2012s [17]. It is carried out in line with the guidelines recommended by the WHO in 2008 as part of the renewal of primary health care [18], the reform of public administration in the DRC and the vision set out in the strategy for strengthening the health system in the DRC [10]. The intermediate level includes the provincial health inspectorates (PHI) with functions focused on control and the provincial health divisions (PHD) with functions oriented more towards socio-technical support for the revitalization and development of health districts [19]. These two reformed structures of the intermediate level of the health system operate under the authority of the provincial government, while maintaining functional links with the structures for the provision of care and other health services at the provincial level and certain hierarchical links with the central level of the Ministry of Health [17]. In order to fully fulfil their missions, the PHDs needed to have competent, motivated executives who met the profile in accordance with the new organic framework. The establishment of the PHDs was thus accompanied by an open and transparent selection process (with the organization of recruitment tests) which resulted in the appointment by decree of the Minister of its facilitators [20, 21, 22].

### Description of intermediate level reform (Provincial Health Division) adopted in 2014-2015

The reform of the intermediate level of the health system in the DRC focused on the duplication of the provincial health inspection into two independent but complementary structures. On the one hand, the Provincial Health Division responsible for the technical supervision of the operational level (health zones) in the implementation of their development plans drawn up in accordance with the directives of the national level and in compliance with the law on decentralization (the function of the Head of the Health Division being under the direct responsibility of the provincial government). On the other hand, the provincial health inspection responsible for the inspection and monitoring of the application of standards and regulations in public health, a structure decentralized in the province but under the direct responsibility of the national government. Despite the fact that health is decentralized in the DRC, the staff of the DPS were recruited by an independent selection committee set up by the Minister of Health and composed of experts from the Ministry of Health (Directorate of Studies and Planning) and delegates from the various technical and financial partners involved in the health sector in the DRC. The recruitment process was led by the National Steering Committee for the Health Sector, in which the Inter-Donor Group for the Health Sector and other ministers of the DRC Government who collaborate with the health sector actively participate. The 7 members of the PHD management team (1 division head and 6 office heads) were recruited on the basis of a national call for tenders, followed by a written test for candidates meeting the criteria and an interview for pre-selected candidates. The other PHD agents were recruited on the basis of an internal call for tenders in each PHD and the analysis of the files by the ad hoc committee [13].

### Justification and objective of the study

More than a decade after the launch of the health system reform implementation process in the DRC and the latter has recently embarked on the path to UHC, very few studies have documented the influence of the reform on the different pillars of the health system. The few studies we consulted focused more on aspects related to the structure of the reform at the intermediate level [18–20], governance and provincial support of the health district [21], financing of the single contract at the PHD level [14] and others on the supervision and performance of service delivery at the intermediate level [22,23]. No study has been interested to date in evaluating the reform in the pillar of human resources management for health at the intermediate level, while according to the WHO they occupy the central place when aiming for universal health coverage [24], path on which the DRC has embarked despite its state of fragility and crisis.

The question is: ‘What is the capacity of the State to carry out a reform of human resources for health at the intermediate level of the health system in a context of fragility and crisis as is the case in the DRC?’

Our study aimed to evaluate the reform concerning the management of human resources of PHD in a context of crisis and fragility. Specifically, it aimed at a dual objective, first to evaluate compliance with the profile in the recruitment of PHD executives and then to identify the factors guaranteeing the retention (loyalty) of the recruited executive staff in order to guarantee the performance of its health system in this particular context.

**Figure 1:**
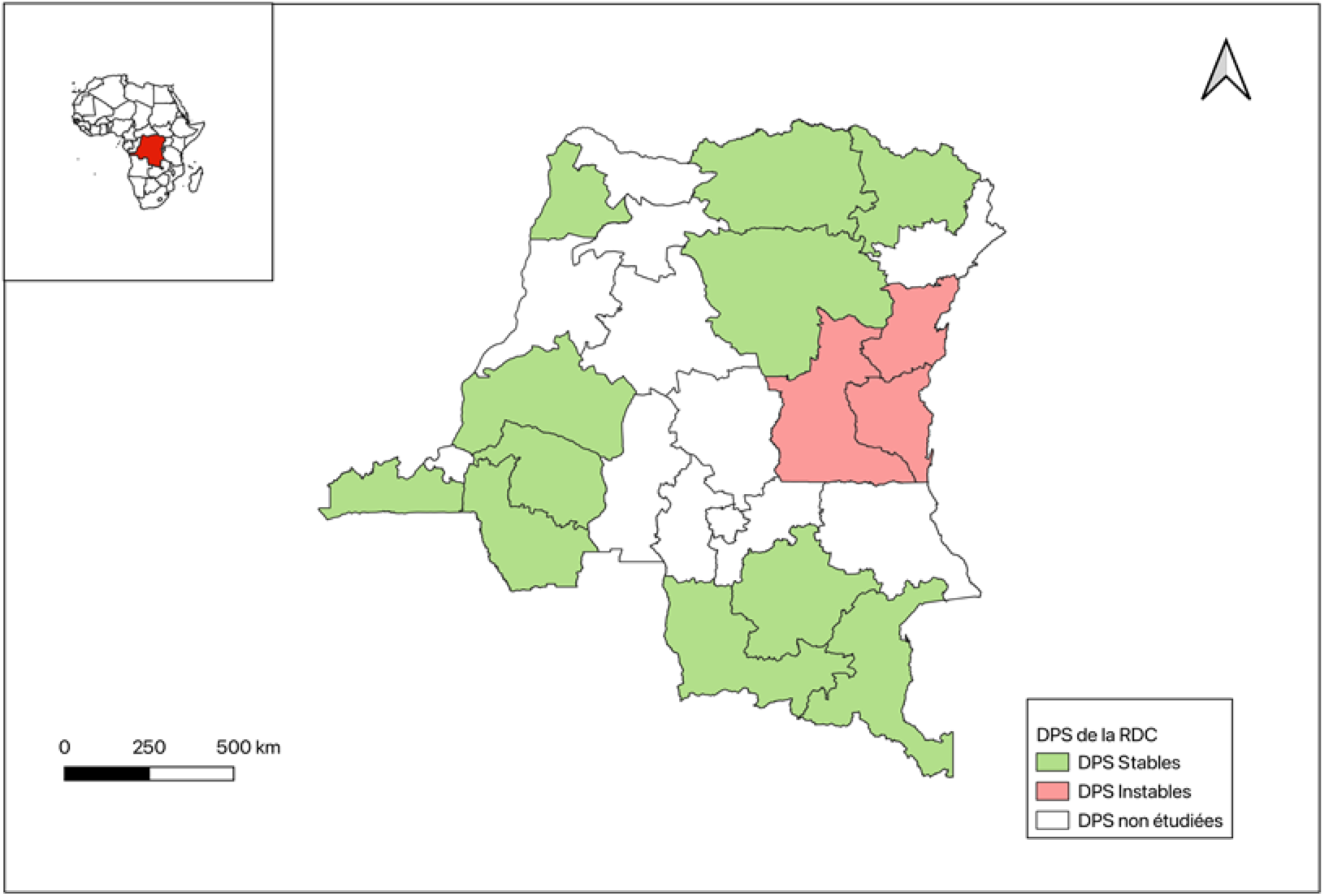
Mapping of provincial health divisions according to stability in the DRC.

## Materials and Methods

### Type and period of study

This was an analytical study of the cohort of staff of the Provincial Health Divisions (PHD) of the DRC and whose sample was constituted by all the agents of these 14 targeted PHD since 16 September 2016, the year of the start of data collection until 13 December 2021.

### Choice of study PHD

The 14 Provincial health divisions concerned are those in which the Directorate of Studies and Planning (DSP) of the Ministry of Public Health of the DRC had organized status missions in relation to the process of reform of the intermediate level of the health system underway in the country. The PHD that met the following two criteria were included in the study: 1) accessibility in relation to the capital city of Kinshasa and 2) the opportunity for funding to organize a field mission in the DPS during the period. The 14 targeted PHD were: Bas-Uélé, Haut-Katanga, Haut-Lomami, Haut-Uélé, Kwango, Kwilu, Kongo-Central, Lualaba, Mai-Ndombe, Maniema, North Kivu, South Kivu, South Ubangi and Tshopo. They represent more than 53.8% of the PHD in the DRC. It should be noted that 3 provinces (North Kivu, South Kivu and Maniema) are considered to be provinces in crisis and unstable following the security situation caused by wars waged by rebels and gangs for decades and which have worsened over the last 5 years.

## Data collection

The data were collected in two stages from all agents of the 14 PHD present during the survey constituting the cohort of this study, first in September 2016 at the implementation of the reform of the intermediate level of the health system and in December 2021, 5 years later. Our sample consisted of the 657 agents present at the start of the survey in 14 PHD out of the26 existing, i.e. 53.8%.

Data were collected directly from PHD agents through a structured questionnaire. They were then supplemented by the files of agents available in each PHD. All agents working in the 14 PHD were included in the study. Excluded were agents made available to the PHI after the reform. Sociodemographic, economic and academic variables were collected from staff in each of the targeted PHD from the managers of the office in charge of human resources. The classification of agents in relation to their level of education was carried out according to defined criteria, in particular the master’s degree and seniority in the management and coordination of health action at the level of the health system, in accordance with the guidelines of the call for tenders launched by the Ministry of Health during the recruitment of PHD management staff.

To calculate the performance level of the PHD, we used the scores obtained at the national level [29]. This performance level was calculated on the basis of a “Benchmarking” score from 27 health indicators commonly collected at the PHD level (indicators of management of killer diseases, indicators of maternal and child health and certain PCA indicators) [29]. A performance score of more than 75% was considered very good.

Management staff recruited in 2016 and still present at the time of the survey in 2021 were considered stable.

We used QGIS software to generate the map of the DR Congo PHD according to their stability.

## Data processing and analysis

Data were entered into an Excel 2016 database and processed with SPSS version 25 software. Quantitative variables were described as means (with SD) or Median (25th percentile – 75th percentile) according to their distribution and categorical qualitative variables as numbers and proportions. The chi-square test was used to compare proportions. The significance threshold was set at 5%.

To describe the specific characteristics of the different PHD, we carried out different comparisons between the PHD depending on the crisis context or not.

As a measure of association, we used relative risks with their 95% confidence intervals. We then constructed multivariate logistic regression models to determine the factors associated with poor PHD performance. Adjusted odds ratios were generated along with their 95% confidence intervals. The error threshold α was set at 5%.

## Ethics approval and consent to participate

This study was approved by the ethics committee of the Catholic University of Bukavu, DR Congo. An informed consent to participate was obtained from all the participants in the study.

## Results

### 1. Characteristics of PHD agents in 2016

Analysis of data on 657 Provincial Health Division workers in 14 provinces of the DRC at the start of the cohort in 2016 reveals key insights, with particular attention paid to the distinction between groups operating in provinces considered unstable and stable (Tab I).

**Tab. 1:**
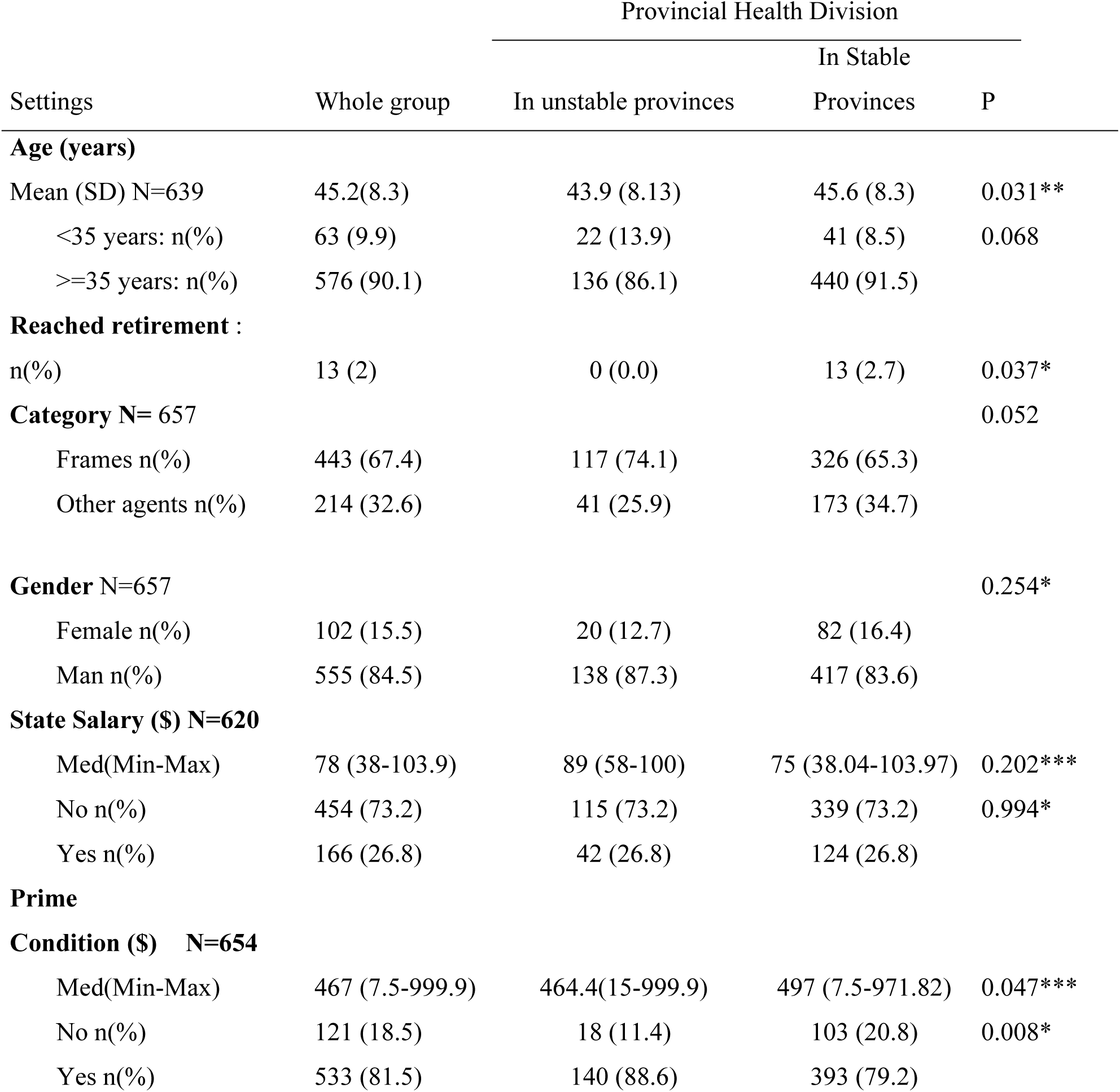
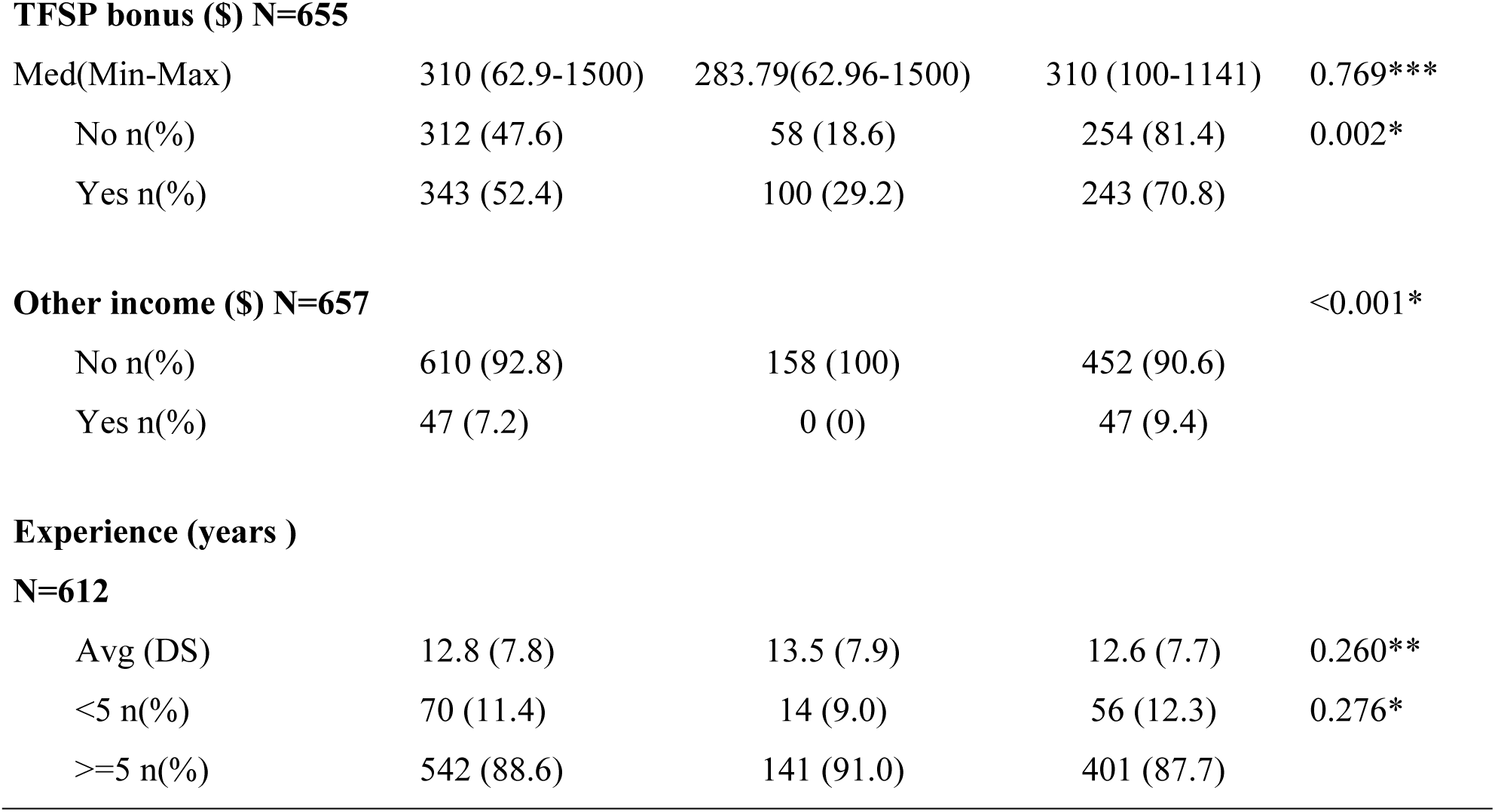
Characteristics of DPS agents in 2016.

**Tab. 2:**
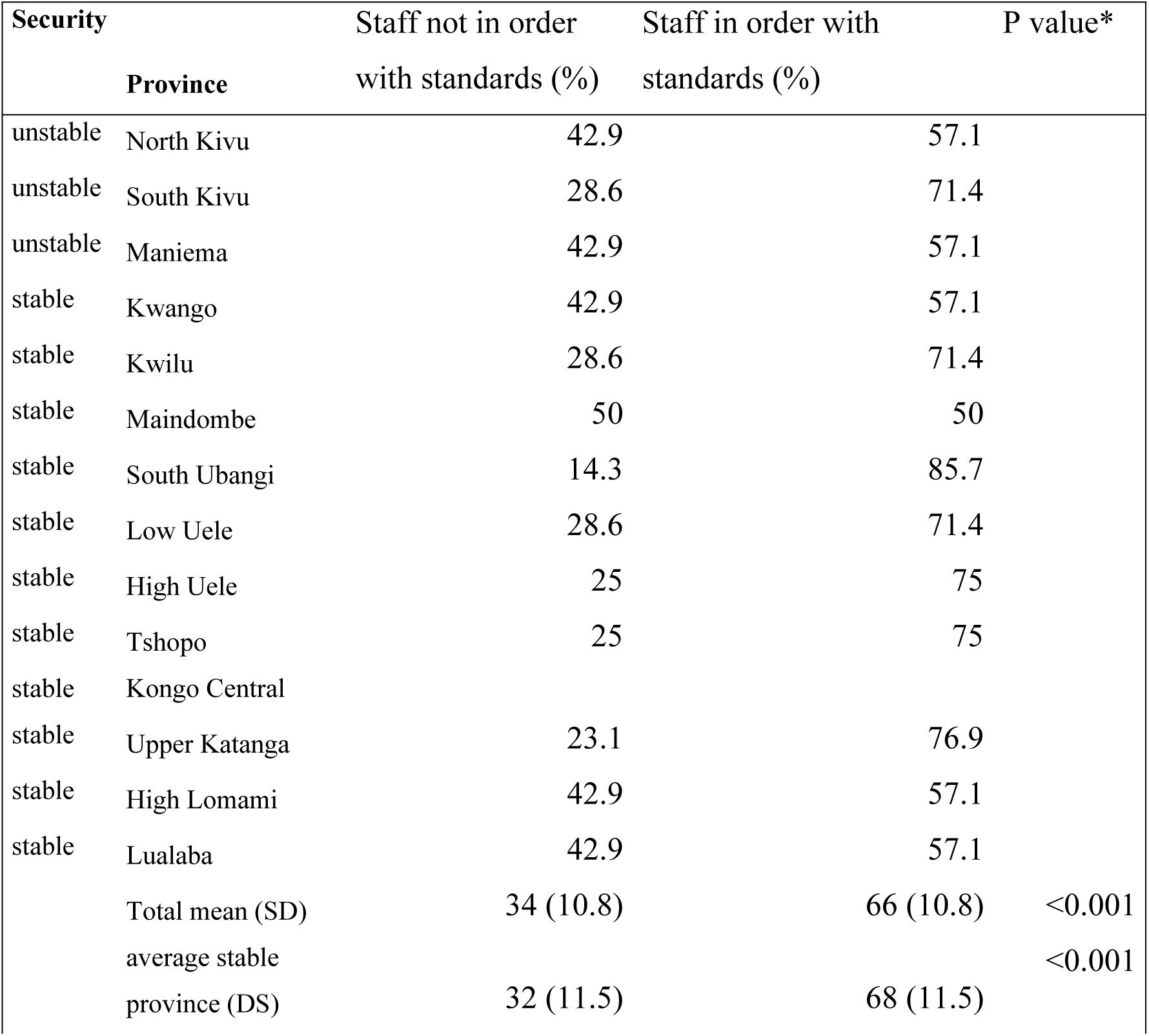
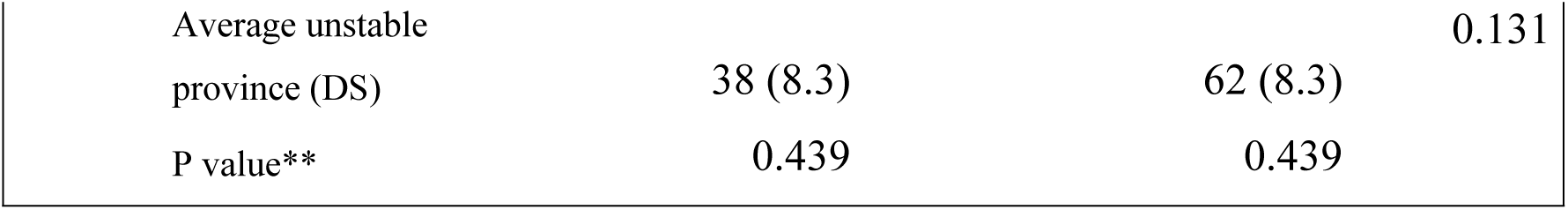
Compliance of recruitment profiles of PHD executive officers with standards in 2016.

| Security | Province | Staff not in order<br>with standards (%) | Staff in order with<br>standards (%) | P value* |
| --- | --- | --- | --- | --- |
| unstable | North Kivu | 42.9 | 57.1 |  |
| unstable | South Kivu | 28.6 | 71.4 |  |
| unstable | Maniema | 42.9 | 57.1 |  |
| stable | Kwango | 42.9 | 57.1 |  |
| stable | Kwilu | 28.6 | 71.4 |  |
| stable | Maindombe | 50 | 50 |  |
| stable | South Ubangi | 14.3 | 85.7 |  |
| stable | Low Uele | 28.6 | 71.4 |  |
| stable | High Uele | 25 | 75 |  |
| stable | Tshopo | 25 | 75 |  |
| stable | Kongo Central |  |  |  |
| stable | Upper Katanga | 23.1 | 76.9 |  |
| stable | High Lomami | 42.9 | 57.1 |  |
| stable | Lualaba | 42.9 | 57.1 |  |
|  | Total mean (SD) | 34 (10.8) | 66 (10.8) | <0.001 |
|  | average stable<br>province (DS) | 32 (11.5) | 68 (11.5) | <0.001 |

|  |  |  |  |
| --- | --- | --- | --- |
| Average unstable province (DS) | 38 (8.3) | 62 (8.3) | 0.131 |
| P value** | 0.439 | 0.439 |  |

The average age of all employees is 45.2 (8.3) years. However, there is a statistically significant variation between the two groups, with an average age of 43.9 years for employees in unstable provinces, while those operating in stable provinces have an average of 45.6 years. ‘Among employees, only 13 (2%) have reached retirement age’ in stable provinces, and the number is zero in unstable provinces.

Not receiving a state bonus offered 2 times less risk [RR=0.5 (95% CI=0.3-0.8), p <0.01] of working in an unstable area compared to an agent who has one and the difference was statistically significant.

Similarly, not receiving a TFSP bonus offered 1.7 times less risk [RR=0.6 (95% CI=0.4-0.8), p <0.01] of working in an unstable area compared to an agent who has one and the difference was statistically significant.

#### Compliance with the standards of the recruitment profile of DPS executives in 2016

In view of the table No. 2, we note that out of all 14 provinces, only 66% of the recruited staff were in compliance with the recruitment standards set by the Ministry of Health against 34% not meeting the standards. We note more agents recruited in compliance with the standards in the stable provinces (68%) than in the unstable ones (62%). This difference is significant (p < 0.001). The province of Sud Ubangi (stable) comes first with staff recruited according to the standards at 85.7% followed by that of Haut Katanga (stable) with 76.9%. Only 50% of the staff in the province of Maindombe (stable) were recruited in accordance with the standards issued.

### 2. Characteristics of DPS agents in 2021 and reasons for absence

In our cohort, 18% of Provincial Health Division (PHD) agents were no longer part of the various DPS in 2021. Of those who remained, 92% kept their initial position, 6% were subject to an internal transfer with a promotion, and 2% were transferred to another internal position without promotion.

The proportions of absent agents by different categories of variables studied show that among agents under 35, 19.4% of agents in this age group are absent in 2021. Those aged 35 and over, 18.3% of agents in this age category are absent. Among those who have reached retirement age, 30.8% are absent in 2021.

In unstable provinces, 22.3% of agents are absent, showing a slightly higher % of absence compared to stable provinces, but without significant difference. For executives, 19.8% are absent in 2021, while other agents present an absence % of 14.6%, without significant difference. Men have an absence % of 19.5% [RR=1.89(1.02-3.48)*p=0.04], showing a statistically higher absence compared to women (10.3%). Among those who do not receive a salary from the State, 17.5% are absent in 2021 with a non-significant difference with those who receive a salary from the State (20.7%). Agents with less than 5 years of experience show an absence rate of 21.7% in 2021 with a non-significant difference with those with 5 years or more of experience (17.6%)(tables 3).

The table 4 compares the “Unstable” and “Stable” groups on several income-related variables. The distribution of state salaries between the “Unstable” and “Stable” groups is almost identical, with approximately 73% of participants in each group not receiving a state salary and 27% receiving one. The statistical test, with a p-value of 0.994, shows that there is no significant difference between the two groups regarding this type of income. State Bonus *(SP*):

There is a significant difference between the two groups for the receipt of the state bonus. Indeed, 88.6% of individuals in the “Unstable” group receive it compared to 79.2% in the “Stable” group. This difference is statistically significant with a p-value of 0.009.

*Technical and Financial Partner (TFP) Bonus:*

Regarding the TFP bonus, 63.3% of agents in the “Unstable” group receive it, compared to 48.9% in the “Stable” group. This difference is also statistically significant, with a p-value of 0.002.

No person in the “Unstable” group reports receiving other income, while 9.4% of agents in the “Stable” group receive it. The difference between these two groups is highly significant, as indicated by the p-value of less than 0.001.

Finally, the median overall income of agents is higher in the “Unstable” group (467) than in the “Stable” group (322). This difference is statistically significant with a p-value of 0.0315(Table 4).

After adjustment with all the factors studied, the factors associated with the absence of agents in the PHD remained the achievement of retirement age [RR (95% CI) = 5.5 (1.2-24.9)] and gender [RR (95% CI) = 3.2 (1.3-7.9)](Supplementary table 1)

No factor studied is associated with the loyalty of PHD executives. The observed trend consists of having a greater risk of stabilizing in PHD for women, executives with more experience (more than 5 years), older ones (more than 35 years), those in stable provinces and those with the PTF-PP bonus, without statistically significant difference (Supplementary Table 2)

The overall performances of the PHD did not show significant differences between 2017 and 2021 (p>0.05). However, we note that some PHD in the stable provinces have decreased their performances (especially the PHD Maindombe, Tshopo and Lualaba). On the other hand, in the 3 unstable provinces, Maniema and North Kivu have fallen, but by only one percent of the score. The comparison between stable and unstable provinces revealed no significant differences in performance (p = 0.349 in 2017 and p = 0.351 in 2021).

Furthermore, staff compliance with the standards showed significant differences with p values <0.001 for the overall score. But this is more observed in stable provinces (p < 0.001) (supplementary table3).

## Discussion

This study evaluates the reform of the health system in its pillar of human resources management for health five years after its effective implementation in the Democratic Republic of Congo, taking into account the particular context of crisis and fragility. Specifically, the study aimed firstly to assess compliance with the profile in the recruitment of PHD executives and secondly to identify the factors guaranteeing the retention (loyalty) of the executive staff recruited in order to guarantee the performance of the Congolese health system.

### 1. Compliance with the recruitment profile criteria for DPS executives in the reform

The results of this study show that in 2016, only 66% of the staff recruited in the 14 PHD met the standards defined by the reform. This observation raises concerns about the effectiveness of the recruitment implemented under the reform, calling into question the ability of the provinces to select staff who met the established standards. However, the reform, in its HRH pillar, aimed to provide the PHD with quality staff, in sufficient numbers and motivated, capable of managing the health system at the intermediate level 19]. It is to achieve this that a participatory process with the organization of recruitment tests was launched and in order to allow each DPS to have competent executives who meet the profile. These results do not show that this process has made it possible to adequately achieve this objective.

We also note that there was a significant difference (p < 0.001) between the level of compliance with recruitment profiles between stable and unstable PHDs (in a state of conflict), the latter only have 62% of personnel who comply with the standards compared to 68% in stable PHDs.

This situation could be explained by the fact that on the one hand, competent executive officers are not motivated to work in these regions in conflict and in a state of insecurity in which they are regularly subjected to threats and attacks by armed gangs. These facts were also reported in a study conducted by Mushagalusa CR at al on “The Profile of Health Personnel in a Context of Instability, a Cross-Sectional Study in 4 Health Zones of South Kivu in Eastern DRC in 2023” [30]. The author had indeed found that agents with a good profile were found more in stable health zones than in those doomed to insecurity for fear of threats. It should be noted that in unstable provinces cases of killings of health executives and other health professionals have been reported especially in the last ten years [31]. The WHO in its 2006 World Health Report had also described how situations of armed conflict impacted on the availability of health personnel [32]. In its report on health in armed conflicts, the International Red Cross had also mentioned similar assertions [31].

What is questionable in our study is to find that even some stable PHD have not also fully recruited staff meeting the required profile. Even a province close to the capital of the country (Maindombe) has recruited only 50% of the staff meeting the set criteria! Indeed, in the context of the DRC, a fragile state whose governance remains weak, our field experience shows that political and geopolitical influences often influence the recruitment process especially in positions of responsibility such as division head and office head. Some provinces are not full of executives with master’s degrees in public health and/or proven experience in health system management but whose “sponsors” are native and politically or security-wise influential are selected and promoted to these positions of responsibility. Similar situations were described in a study cited above at the level of recruitment of health care providers at the operational level of the health system [30]. Given the growing number of public health science training institutions at the master’s level in the DRC, we dare to believe that it is not the skills that were lacking in the provinces but rather the political will to respect the required profile for the reasons mentioned above. Unfortunately, this practice has prevented the PHD from being managed by executives who would enable them to achieve their objectives in terms of coordination and support of health zones for the improvement of the health status of the population. It should also be noted that the recruitment process was decentralized and conducted locally in the provinces with the support of experts from the Ministry of Health and the TFPs; which would have minimized too many local political influences. The unstable PHD have therefore not only been subject to these political influences but also because agents with good profiles were not willing to work there following the states of insecurity.

This observation calls into question the capacity of the provinces alone to lead the process of recruiting human resources for health in a context of instability and fragility and demonstrates the need to involve the national level and technical and financial partners in this process for its success.

### 2. Characteristics of DPS personnel in 2021, i.e. after 5 years of the reform

The results of this survey show in its table n° 3 that in the cohort studied, 18% of PHD agents present at the start of the survey in 2016 were absent at the end of the survey in 2021. These data highlight the diversity of reasons for absence within the DPS. Among the causes of absence recorded, the death of 11% of agents is a tragic reality that affects the workforce. In addition, 16% of agents were promoted to other functions within the health system, illustrating career opportunities within the organization. The desertion or dismissal of 16% of agents raises important questions about the underlying factors and challenges related to staff retention. Furthermore, the availability of 48% of agents, who now work within non-governmental organizations (NGOs) or partners of the Ministry of Public Health (MSP), reflects the mobility of health personnel within the health sector and cooperation with external partners.

In the unstable provinces, 22.3% of agents were absent, showing a slightly higher absence rate compared to the stable provinces in which this rate is lower (16.6%). This could be explained by the context of fragility and conflict suffered by the populations of the 3 unstable provinces. Thus, some agents who cannot resist the effects of insecurity (threats, thefts, looting, etc.) become discouraged and look for work elsewhere. Other agents agree to go and work in the unstable provinces when they have not found a job elsewhere; once they find opportunities outside these provinces, they do not hesitate to leave. This situation corroborates with that documented by other authors such as WHO 2022 [33]., Witter S and Pavignani B 2018 [34]., Mushagalusa CR et al, 2023 [30]. who also found that the state of conflict and fragility hindered the proper functioning of the health system, especially by depriving it of adequate and competent manpower to achieve its objectives. In this same context, a study monitored in 2022, nearly 2,000 attacks against health structures and care providers in areas affected by conflict were recorded in more than 30 countries. 159 attacks of this type were recorded in the DRC, killing at least 11 health workers [35] [36] This crisis context could have a significant part in the absences observed in unstable provinces. What is still interesting is to note that most of those leaving the DPS continue to work in the health system (partner NGOs, PTFs, national level, etc.).

### 3. On the remuneration (income) of DPS management staff during the reform

According to the results of this study, we note that 5 years after the implementation of the reform of the intermediate level of the health system in the DRC, only 27% of the agents of the 14 PHD under study received the salary of the Congolese State against 73% did not receive one. This situation is similar both in the stable provinces and in those unstable for this type of income (p = 0.994). This is worrying if we consider that it is the structure of the intermediate level that coordinates the operational level of the health system and ensures its junction with the national level. Indeed, the agents of the peripheral level must address and bring their grievances to the intermediate level to plead in their favor at the national level among other things for the alignment with the salaries of the State. It then becomes almost impossible to carry out this mission by this level itself overflowing with more than 70% of the non-salaried agents. This corroborates with the results found by Mushagalusa CR et al in a 2017 study on the profile of operational level staff in 4 rural health zones in the DRC which found that 95% of health workers did not receive a state salary [30].

As for the bonuses granted by the State, our results show a statistically significant difference (p of 0.009) between the two groups; 88.61% of agents in unstable provinces receive it against 79.23% in stable provinces. Can we think that agents in unstable provinces are more advantageous given their level of exposure to security conditions?

These results are different from those of the study by Mushagalusa CR et al who found that only 34% of the staff of the 4 health zones of the province of South Kivu receive the State bonus and without significant difference between the unstable zone and the stable one. This difference between the two studies could be explained by the fact that the first was done on a small scale unlike the second.

As for the bonuses granted by technical and financial partners (TFPs), our data clearly show a significant difference between the two groups (p=0.002). Indeed, 63.3% of agents in unstable provinces receive this bonus compared to 48.9% in unstable provinces. This difference can be attributed to the greater presence of TFPs in conflict provinces in which most NGOs carry out emergency actions, which is not the case in stable provinces.

As for the overall income of PHD agents, the results of the study show a significantly higher (p = 0.0315) median overall income of agents in the “unstable provinces” group ($467) than in the “stable provinces” group ($322). The difference observed between the unstable and stable groups could be attributed to the fact that agents in the unstable group more frequently benefit from the bonus from technical and financial support partners (TFPs). This situation could be explained by the emergency context, where unstable agents, often on the front line, are more likely to receive this type of support to compensate for the precariousness of their situation.

### 4. Factors for stabilizing or retaining staff in PHD

Our study did not find among the factors studied those associated with the stability (loyalty) of PHD staff as indicated in Table No. 6, it nevertheless noted a tendency of risk of stabilization in PHD for women, executives who have more experience (more than 5 years), the oldest (more than 35 years), those working in stable provinces and those with the technical and financial partners bonus (PTF), without statistically significant difference. Our experience in the field actually shows that older executives, those with more than 5 years of experience as well as women are in most cases natives and residents in the area with their families or who have decided to settle permanently in an area and almost no longer want to move because they have either invested or carried out other lucrative activities (teaching, private clinics, etc.).

For those in unstable and conflict-ridden provinces, they are also already accustomed to the conditions of insecurity, where they know the actors in place and their modus operandi, as well as the concessions or “arrangements” to be made in order to have peace.

However, it should also be noted that the permanent presence of humanitarian NGOs and technical and financial partners in the provinces in conflict may justify a tendency of some PHD executives to want to stay in these areas following the bonuses and other benefits they offer to motivate agents to continue to take care of the health of the population. Table No. 4 of this study clearly shows that agents in unstable provinces benefited more from bonuses from TFPs than those in stable provinces.

According to a study conducted in 2018 in Côte d’Ivoire on the motivation of health personnel and students to work in underserved areas, it emerged that the combination of incentive measures including the existence of essential amenities such as drinking water and electricity, the provision of on-call housing plus a bonus of 20% of the current salary constituted the most relevant incentive packages for all respondents [37].

### 5. On the performance of DPS during the reform based on compliance with the profiles of recruited agents

This study reveals interesting information that deserves in-depth analysis; indeed, we realize that the average overall performance of the 14 PHD remained the same 5 years after the reform, i.e. between 2017 and 2021 (72% versus 71%). The analysis of provincial performances reveals a worrying situation, characterized by high percentages of staff not respecting the standards defined by the reform, reaching on average 34%. Some provinces, such as Kwango, Kwilu, Kongo Central and Haut Lomami, have managed to significantly improve their performance. These positive results suggest a successful implementation of the reforms, perhaps through more rigorous recruitment processes and effective training programs. However, other provinces, such as Maindombe, Tshopo and Lualaba have experienced notable declines, highlighting specific challenges in the implementation of the reform, particularly in terms of recruiting and retaining staff who meet standards.

However, other provinces, such as Maindombe and Haut Lomami, have seen notable declines, highlighting specific challenges in implementing the reform, particularly in recruiting and retaining staff who meet standards.

Regarding provincial stability, it is interesting to note that performance remains stable in provinces considered unstable, such as North and South Kivu. Despite their instability, these provinces have managed to maintain good performance, probably due to substantial support from partners in the context of chronic instability as shown in Table 4. In contrast, the province of Maniema, also unstable, presents a weak performance, possibly due to its isolation and the lack of support similar to that of the other two unstable provinces.

**Table No. 3:** Reasons for absence within the Provincial Health Division in 2021.

| Becoming agents who left the PHD between 2016-2021 | n(%) |
| --- | --- |
| Death | 12 (10.62) |
| Promotion to another position in the health system | 18 (15.93) |
| Desertion or revocation | 18 (15.93) |
| Availability (work in NGOs or MSP partners) | 54 (47.79) |
| Others | 11 (9.73) |

**Table No. 4.**
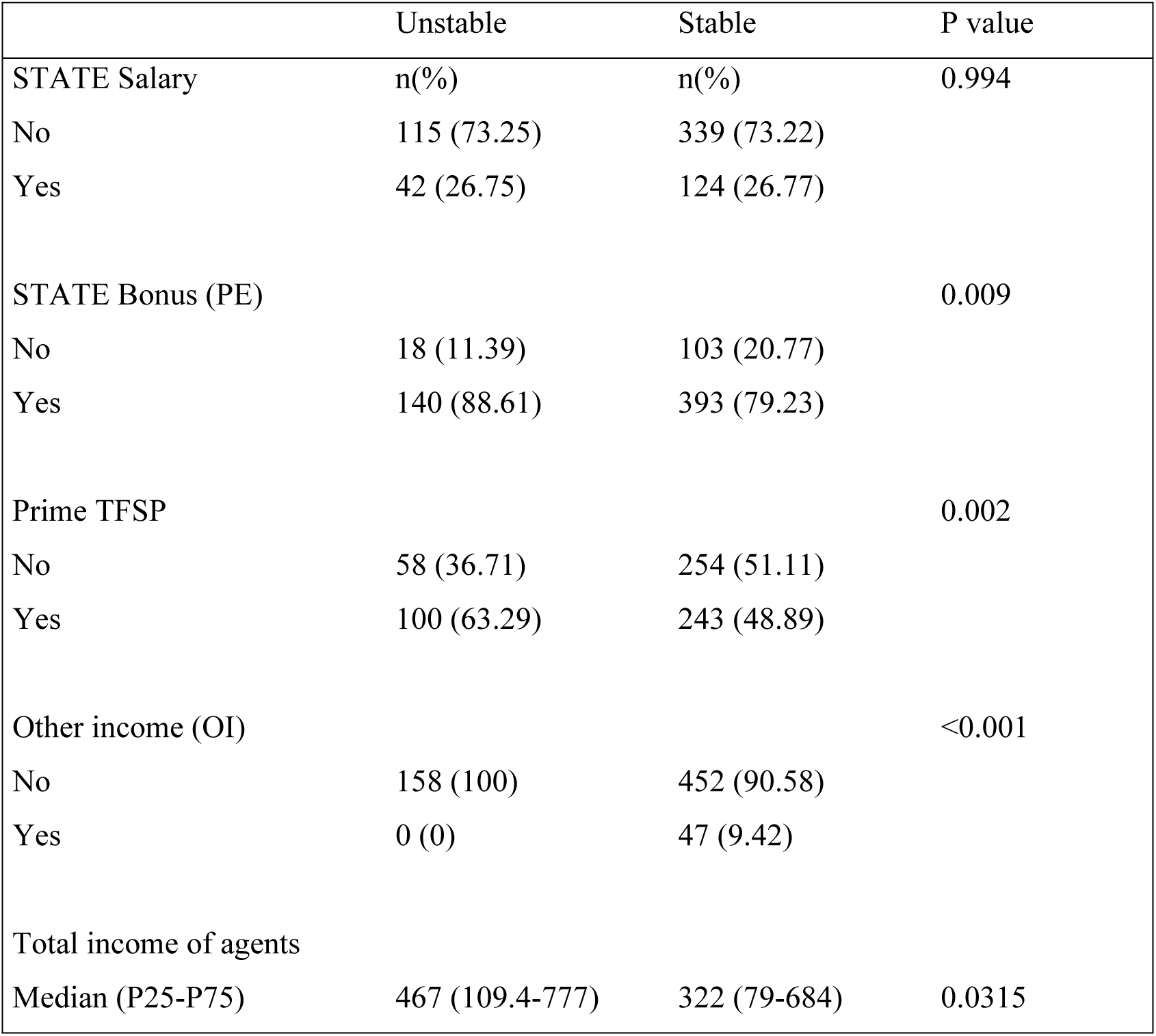
Income of PHD agents according to the level of stability of the provinces.

Stable provinces maintained a relatively constant average, ranging from 71% to 70% before and after the implementation of the reform. This indicates some consistency in personnel management in accordance with established standards, highlighting the importance of stability in maintaining satisfactory performance.

## Methodological limitations

The results of this study concern only 14 out of the 26 PHD of the country, it is possible that other realities are found in the PHD not included. Fortunately, the PHD surveyed are representative of the 11 former provinces and the sister PHD have almost the same characteristics as the mother provinces at least concerning human resources.

In addition, we did not seek to know at what time (year) each agent left the cohort, in fact it was practically difficult to calculate the cumulative incidence given that the agents who abandoned work hardly write and their abandonments are noted or declared 6 months or years later. Most seek to keep a foot in the door with the hope that the State will one day improve the living conditions of its workers.

## Conclusion

The recruitment process launched at the beginning of the reform and which aimed to provide the PHD offices with competent agents who met the required profiles was not fully followed. This decentralized recruitment at the local (provincial) level could obviously not escape the influences of the realities of the current socio-political and security context of the country. This is more felt in the unstable provinces in which the level of governance is weakened following the presence of armed groups. This observation calls into question the capacity of the provinces alone to lead the recruitment process of human resources for health in a context of instability and fragility.

There is a problem of retention or stabilization of PHD staff leading to frequent departures of executives due to the absence of an incentive policy including financial and non-financial incentives at the country level; this instability of agents has been reinforced by the security situation in unstable provinces. The non-alignment of state salaries for the vast majority of PHD agents is a glaring weakness throughout the country requiring a courageous decision; the same is true for the security of personnel which are all factors guaranteeing the loyalty of agents. The presence of technical and financial partners fortunately contributes to maintaining a minimum of stability of personnel in unstable provinces and their efforts need to be supplemented and supported by the State.

The gap in provincial performance suggests the need for a thorough evaluation of staff recruitment, training, and compliance processes, with specific adjustments to local realities to maximize the impact of reforms.

This study shows that even in a context of crisis and fragile state, health system reform is possible. It is possible to organize staff recruitment through a selection process independent of the political authorities of the Ministry of Health and supported by the technical services of the Ministry and partners. The involvement of technical and financial partners (TFPs) in the recruitment process helped the Ministry of Health to minimize political influence in the recruitment of middle-level executives. Experience and the presence of a financial bonus are motivating factors for staff stability.

## Data Availability

The minimal data set is available on a reasonable request to the corresponding author

## Acknowledgments

We sincerely thank all the people who contributed to this work. The authors sincerely thank the team of the General Secretariat for Health of the DRC and its directorates, all 14 Heads of provincial health divisions and their management teams for their collaboration during this study.

## Authors’ contributions

CRM, BBG, DGM and MMA: designed the study protocol, supervised data collection and edited the final manuscript.

ANG, DGM and CCM contributed to data collection in the 4 targeted health districts.

PM and MTA: contributed to data analysis and writing of the work.

Other authors interpreted data and contributed to the enrichment of the manuscript. All authors reviewed and approved the final manuscript.

## Financing

Not applicable.

## Availability of data and materials

The datasets used and/or analyzed during this study are available from the corresponding author upon reasonable request.

## Consent to publication

Not applicable.

## Abbreviations

Average(SD)=Mean (Standard Deviation);

Med(P25-P75)=Median (25th percentile – 75th percentile);

*Chi-square;

** Student’s T;

***Man Whitney;

TFSP=Technical and Financial Support Partners; $=US Dollar

NGO = Non-governmental organization; MPH = Ministry of Public Health

